# Changes in the prevalence of the common risk factors for non-communicable diseases in Uganda between 2014 and 2023: Informed by nationally representative cross-sectional surveys

**DOI:** 10.1101/2024.09.04.24313080

**Authors:** Ronald Kusolo, Gerald Mutungi, Mary Mbuliro, Richard Kajjura, Ronald Wesonga, Silver K. Bahendeka, David Guwatudde

## Abstract

**Background:** Non-communicable diseases (NCDs) remain the biggest contributor to global mortality. Although communicable diseases still contribute highest to mortality in the African region, the proportion contributed by NCDs continues to rise. An important way to control NCDs is to focus on reducing the prevalence of the common NCD risk factors. As such, monitoring changes in the prevalence of these risk factors is essential for better NCD prevention planning. Uganda conducted its first nationally representative NCD risk factor survey in 2014, and a second in 2023. We analyzed the prevalence of the common NCD risk factors using data from the two surveys to identify which risk factors changed between 2014 and 2023.

**Methods:** Both surveys drew countrywide samples stratified by the four regions of the country, and by rural-urban locations. The World Health Organization’s STEPs tool was used to collect data on demographic and behavioral characteristics, and physical and biochemical measurements. We defined and calculated weighted prevalence of the following common NCD risk factors: high blood pressure, high blood glucose, overweight and obesity, current use of alcohol, current tobacco use, inadequate consumption of fruits and vegetables, inadequate physical activity, and sedentariness.

**Results:** The 2014 survey enrolled a total of 3906 participants, whereas that of 2023 enrolled 3694 participants. The NCD risk factor prevalences that increased significantly were: high blood glucose from 1.3% in 2014, to 2.9% in 2023 (p< 0.001); overweight and obesity from 19.4% in 2014 to 24.1% in 2023 (p< 0.001); current alcohol consumption from 28.5% in 2014, to 31.1% in 2023 (p=0.013); and sedentariness from 26.6% in 2014, to 31.9% in 2023 (p< 0.001). It is only the prevalence of current smoke tobacco use that decreased significantly from 9.6% in 2014, to 8.3% in 2023 (p= 0.046). Nationally, no significant changes were observed in the prevalence of high blood pressure, inadequate consumption of fruits and vegetables, and inadequate physical activity.

**Conclusions:** Most of the common NCD risk factors increased between 2014 and 2023. There is an agent need for the various stakeholders in Uganda to implement interventions targeting reduction in the prevalence of NCD risk factors to prevent the currently increasing burden of NCDs and associated mortality.

## INTRODUCTION

Non-communicable diseases (NCDs), mainly cardiovascular diseases (heart disease and stroke), cancer, diabetes, chronic respiratory diseases, and poor mental health, kill approximately 41 million people each year, equivalent to 74% of all deaths globally. Of these, 17 million are premature deaths that occur before the age of 70 years; and 86% of these occur in low- and middle-income countries (LMIC) [1]. Thus NCDs are the greatest contributor to global mortality.

Although at a lower rate, the rates of NCD associated-mortality are increasing in the sub-Saharan Africa (SSA) region. Currently, the proportion of mortality in the African region due to NCDs ranges from 36% to 88%. Cardiovascular diseases, cancers, diabetes mellitus and chronic respiratory diseases account for over 70% of NCD-related mortality [2].

An important way to control NCDs is to focus on reducing the prevalence of risk factors associated with these diseases [1]. Surveillance, or regular monitoring of the common risk factors for NCDs is essential in public health to inform decision making for prevention and control of NCDs. The common NCD risk factors for include behavioral risk factors including physical inactivity, alcohol consumption, tobacco use, low consumption of fruits and vegetables, and sedentariness; and physiological/ metabolic risk factors that include overweight and obesity, high blood pressure, hyperglycemia (high blood glucose levels), and hyperlipidemia (high levels of fat in the blood) [1]. A number of LMICs have now conducted nationally representative population based surveys that collect data on the common risk factors for NCDs, mostly using the WHO STEPS methodology [3]. Findings from these surveys can be used not only to monitor the prevalence of the common NCD risk factors, but to also evaluate the effectiveness of any interventions that may have been implemented.

Uganda conducted its first nationally representative NCD risk factor survey in 2014 [4, 5], and a second recently in 2023. Since 2014 Uganda’s Ministry of Health has spearheaded a number of public health preventive initiatives aimed at reducing the general public’s risk for NCDs. But it is not known if there have been improvements in the prevalence of the common NCD risk factors. Of interest is to monitor changes in the risk factors for better NCD prevention planning. We analyzed the prevalence of the common NCD risk factors using data from the Uganda NCD risk factor survey conducted in 2014, and that conducted in 2023. The primary objective of our analysis was to assess any changes, and to identify which NCD risk factors changed between 2014 and 2023.

## MATERIALS AND METHODS

### Study Design

A cross-sectional study design was used to conduct both the 2014 and the 2023 surveys. The 2014 survey was conducted between March and July 2014, whereas the 2023 survey was conducted between February and April 2023.

### Sample Size

The sample size for the survey was calculated assuming a 50% prevalence of the risk factors (*P*= 0.5, which is the recommended value for unknown prevalence, and gives the most conservative sample size), a 5% level of significance (*α*=0.05), a margin of error of 0.05 (*δ*=0.05); an expected response rate of 80%, six age-sex categories (18-29, 30-45, 45-69), and a 1.5 design effect. Using these parameters, the sample size calculation was made using the WHO STEPS sample size calculator [6]. The calculated sample size was a total of 4322.

### Study Population & sampling

Uganda currently has a total population of approximately 45.8 million [7], of which approximately 45% are adults aged 18 years or older. Both surveys covered the whole country, and three stage stratified sampling was used as described below. We used population enumeration areas (EAs) information that was provided by the Uganda Bureau of Statistics (UBOS). UBOS demarcated the whole country into a total of 78,691 EAs that they used for the Uganda Population and Housing Census of 2014. The strata were regions stratified by urban-rural locations, that is, regions (Northern, Western, Central & Eastern), then by urban and rural location within each region, generating a total of 8 strata. The first stage of selecting the study sample was a random sample of 350 enumeration areas (EA) in 2014, and 310 in 2023 from the 8 strata using proportion to size sampling (PPS), as the first stage of sampling. Research Assistants (RA) that had received a five-day training were then dispatched throughout the country to list all households within the sampled EAs. The listing was used to conduct the second stage of sample selection by randomly selecting 14 households within each EA.

A different team (Interviewers), who had also received a five-day training on procedures and administration of the STEPs tool was then dispatched throughout the country. They started by enumerating all eligible members the sampled households and recorded these in an electronic tablet. The final third stage of sample selection involved randomly selecting one eligible household member within each household, which was conducted by the study Interviewers using an android tablet that was pre-set to randomly select one individual from the list of household members for inclusion in the survey, leading to a total sample of 4900 2014, and 4322 in 2023. Sample selection was conducted without replacement, thus there was no replacement of the android tablet pre-selected household member to prevent potential selection bias.

Eligible participants were adults aged between 18-69 years of age, had been members of their household for at least six months preceding the date of the survey, of sound mind, and able to give written informed consent. Household members with the following characteristics were excluded: unable to stand without support (e.g., after limb amputation without a prosthesis), clearly under the influence of alcohol or other drug(s) abuse, and moribund and obtundent.

### Measurements

Both surveys used the World Health Organization’s (WHO) STEPwise approach to surveillance, a standardized method of analyzing risk factors for NCDs [3]. The STEPwise approach is a sequential process starting with collecting data on key risk factors using a questionnaire (STEP 1), followed by physical measurements (STEP 2), and finally collection of biological samples for biochemical assessments (STEP 3).

In STEP 1, data was collected on demographic and behavioral information using the STEPS questionnaire [8]. This step collected information on the demographic and social characteristics (e.g. age, sex, level of education, employment, income, etc); behavioral characteristics (e.g. tobacco use, alcohol consumption, fruit and vegetable consumption, physical inactivity, etc). It also included information on health history like history of raised blood pressure, raised blood glucose, raised blood cholesterol, cardiovascular diseases, lifestyle advice and cervical cancer screening for women respondents, household energy use, mental health (depression) and impact of the COVID-19 pandemic on behavioral risk factors. STEP 2 involved making the physical measurements including height, weight, girth (waist, hip), blood pressure and pulse, which were made immediately after administering the questionnaire. Three blood pressure readings were taken 3–5 minutes apart. After administering the questionnaire and making the physical measurements, Interviewers requested participants to converge at pre-arranged location the following morning so that a blood and urine sample could be obtained to conduct the biochemical measurements. Participants were requested to fast from food and drinks for at least 8 h overnight, and not to indulge in exercise or smoking in the morning prior to the measurements as described in more details elsewhere [9].

STEP 3 was conducted on day two and involved performing biochemical measurements using point of care devices to measure fasting plasma glucose. blood glucose, blood lipids (total cholesterol and High-Density Lipoprotein cholesterol), and spot urine sodium, creatinine and cotinine testing.

### Ethics

The 2023 the survey was approved by the Institutional Review Committee of Nsambya Hospital, Kampala, Uganda (Approval #: SFHN-2022-43), and the 2014 survey was approved by the same IRB (Approval #: IRC/PRJ/11/13/031). Written informed consent was obtained from eligible subjects before enrollment in the study. Participants with at least two systolic blood pressure readings of at least 121 mm Hg, and/or diastolic blood pressure of at least 81mm Hg, and/or with fasting plasma glucose of at least 6.1 mmol/L, and were not already on treatment for hypertension and/or diabetes, were advised to as soon as possible report to the nearest government owned health facility for further evaluation.

### Data management & Analysis

#### Data management

The STEPS data collection tool was programmed using Open Data Kit (ODK), an open-source data collection platform. All data was collected through a password protected tablet to ensure confidentiality and encrypted during transmission from the Interviewers to the server to protect data from unauthorized access. Data quality indicators were monitored through a real-time dashboard by the data manager. All the identified quality issues were discussed with the field team for timely resolution. The final cleaned datasets were then merged and locked for analysis.

#### Statistical Analysis Methods

The response rate was calculated as the proportion of eligible household members who consented to participate in the survey, out of the total number of sampled households. Descriptive statistics are reported as means with corresponding standard deviations for the study continuous variables. Categorical variables, including sex, urban-rural status and region of residence, level of education, marital status, and employment status are reported as frequencies and proportions.

The prevalence of the common NCD risk factors are reported by region, and by urban-rural location (urbancity), comparing their prevalence between the 2014 and the 2023 surveys. Below is a detailed description of how the prevalence of the risk factors were calculated.

The Behavioral Risk Factors included:

1. The prevalence of current alcohol use was calculated as the percentage of participants reporting to had consumed any amount and type of alcohol in the past 30 days preceding the day of the interview.
2. The prevalence of current tobacco use was calculated using three different indicators.

The first was prevalence of current use of all forms of tobacco, calculated as the percentage of participants reporting to had used smoke and/or smokeless tobacco in the past 30 days, including smoking, chewing, and/or sniffing. The second indicator was prevalence of current smoke tobacco use, calculated as the percentage of participants reporting to had used smoke tobacco in the past 30 days. The third indicator was the percentage of participants who tested positive to the Cotinine/Nicotine test (COT200).

1. iii) The prevalence of inadequate consumption of fruits and vegetables was calculated as the percentage of participants reporting to had consumed an average of less than five servings of fruits and/or vegetables combined per day, over the past seven days preceding the day of interview.
2. iv) The prevalence of sedentariness was calculated as the percentage of participants reporting an average of more than four hours of awake time, seated, reclined or lying posture. The cut-off of four hours was based on a study that showed that more than four hours of sedentary increased the risk of all-cause, cardiovascular and cancer mortality, and incident type 2 diabetes [10].
3. v) The prevalence of inadequate physical activity was calculated as the percentage of participants reporting less than 150 minutes per week of moderate intensity physical activity, and/or less than 75 minutes per week of vigorous intensity physical activity, per recommendations by the World Health Organization (WHO) [11].

The Physiological/ Metabolic Risk Factors included:

1. The prevalence of high blood pressure calculated as the percentage of participants with an average two readings of systolic blood pressure ≥ 140, and/or an average of two readings of diastolic blood pressure ≥ 90 mmHg [12, 13], and/or reporting to be currently on hypertension treatment.
2. The prevalence of high blood sugar was calculated as the percentage of participants with a fasting plasma glucose reading of > 7.0 millimoles per liter (mmol/L), or reporting to be currently on anti-diabetes treatment [14].
3. The prevalence of overweight and obesity was calculated as a Body Mass Index greater than 25. BMI for a participant was calculated as weight in kilograms per squared meter (kg/m^2^) [15].

We report weighted prevalence estimates using the study sampling weights. Three distinct weight levels were applied in calculating the prevalence of the above risk factors. Step 1 weights were used for the self-reported behavioral risk factors including: prevalence of current alcohol use, prevalence of current tobacco use (except cotinine/ nicotine test), prevalence of inadequate consumption of fruits and vegetables, prevalence of sedentariness, and prevalence of inadequate physical activity. Step 2 weights were applied in calculating the prevalence of physiological/ metabolic NCD risk factors, including prevalence of high blood pressure, prevalence of high blood sugar, and the prevalence of overweight and obesity.

Step 3 weights were applied in calculating prevalence of current tobacco use as measured by Cotinine/Nicotine detection (COT200).

Chi-square tests were applied to assess statistical differences in the prevalence of the NCD risk factors between the 2014, and the 2023 surveys, with a significance level at 5%. Data analysis was conducted using the STATA software version 15.

## RESULTS

### Response rates in the 2023 Survey

A total of 4340 households were approached to participate in the 2023 NCD Risk Factors STEPS survey, from which one randomly selected eligible adult was requested to consent to participate in the survey. A total of 3694 (1426 men; 2268 women) consented to participate, giving a response rate of 85.1%. The distribution of response rates by region and urbanity are shown in Table 1. For the 2014 survey, out of the 4900 nationwide sampled subjects, 3987 consented to participate in the survey, giving a response rate of 81.4% [4, 5].

**Table 1:** Distribution of Response Rate by Region.

| Region & urbanity |  | Number of<br>Households selected | Number of<br>Households Realized | Response Rate<br>(%) |
| --- | --- | --- | --- | --- |
| Central | Urban | 476 | 357 | 75.0 |
|  | Rural | 616 | 547 | 88.8 |
| Eastern | Urban | 210 | 194 | 92.3 |
|  | Rural | 1008 | 897 | 89.0 |
| Northern | Urban | 126 | 104 | 82.5 |
|  | Rural | 826 | 706 | 85.5 |
| Western | Urban | 224 | 186 | 83.0 |
|  | Rural | 854 | 742 | 86.9 |
| **Total** | - | **4,340** | **3,694** | **85.1** |

### Characteristics of the participants

Of the 3694 participants in the 2023 NCD Risk Factors STEPS survey, 2268 (61.4%) were female, 2,892 (78.3%) were rural residents, 1,560 (42.2%) were aged between 40 years and above, and 968 (26.2%) had attained at least primary school education. By region of residence, 810 (21.9%) were from the northern region, 904 (24.5%) from central, 1,052 (28.5%) from eastern, and 928 (25.1%) from the western region. The average age of participants was 38.3 (SD=13.8). A summary of selected characteristics of the participants is presented in Table 2. Characteristics of participants in the 2014 survey have previously been reported elsewhere [4, 5].

**Table 2:** Participant characteristics.

| Characteristic |  | -n- | (%) |
| --- | --- | --- | --- |
| All participants | - | 3,694 | 100% |
| Sex: | Males | 1,426 | 38.6 |
|  | Females | 2,268 | 61.4 |
| Urban-rural: | Rural | 2,892 | 78.3 |
|  | Urban | 802 | 21.7 |
| Region: | Northern | 810 | 21.9 |
|  | Central | 904 | 24.5 |
|  | Eastern | 1,052 | 28.5 |
|  | Western | 928 | 25.1 |
| Age in years: | 18-19 | 186 | 5.0 |
|  | 20-29 | 1,033 | 28.0 |
|  | 30-39 | 915 | 24.8 |
|  | ≥40 | 1,560 | 42.2 |
|  | Mean ± SD | 3,694 | 38.3 ±13.8 |
| Level of education: | None | 2,122 | 57.4 |
|  | Primary sch | 968 | 26.2 |
|  | Secondary sch | 463 | 12.5 |
|  | University or higher | 140 | 3.9 |
|  | Not stated | 1 | 0 |
| Marital status: | Never married | 455 | 12.3 |
|  | Currently married | 2,033 | 55.0 |
|  | Separated | 414 | 11.2 |
|  | Divorced | 38 | 1.0 |
|  | Widowed | 268 | 7.3 |
|  | Cohabiting | 485 | 13.1 |
|  | Refused | 1 | 0 |
| Employment status: | Employed | 2941 | 79.6 |
|  | Non-paid | 177 | 4.8 |
|  | Student | 64 | 1.7 |
|  | Unemployed | 511 | 13.8 |
|  | Refused | 1 | 0 |

### Prevalence of the common NCD risk factors

Our analysis showed no overall significant change in the prevalence of high blood pressure between the 2014 survey at 24.3%, and that in the 2023 survey at 24.5% (p=0.838); neither were there significant changes between the two surveys in the four regions of the country (Table 3), nor by urbancity (Table 4).

**Table 3:** Prevalence of the common NCD Risk Factors by Region.

| Risk factor | Northern |  | Central |  | Eastern |  | Western |  | Overall |  |
| --- | --- | --- | --- | --- | --- | --- | --- | --- | --- | --- |
|  | 2014 | 2023 | 2014 | 2023 | 2014 | 2023 | 2014 | 2023 | 2014 | 2023 |
| High blood pressure | 19.1% | 19.6% | 27.7% | 27.5% | 26.2% | 27.3% | 23.3% | 21.6% | 24.3% | 24.5% |
|  | 0.801 |  | 0.918 |  | 0.577 |  | 0.377 |  | 0.838 |  |
| High blood glucose | 1.3% | 1.5% | 1.5% | 4.2% | 0.9% | 2.7% | 1.4% | 2.8% | 1.3% | 2.9% |
|  | 0.735 |  | <0.001 |  | 0.003 |  | 0.034 |  | <0.001 |  |
| Overweight and obesity | 8.3% | 14.6% | 30.2% | 36.8% | 16.9% | 18.9% | 19.2% | 23.8% | 19.4% | 23.9% |
|  | <0.001 |  | 0.001 |  | 0.242 |  | 0.015 |  | <0.001 |  |
| Current alcohol use | 36.9% | 41.1% | 28.9% | 26.1% | 20.8% | 26.4% | 28.2% | 34.8% | 28.5% | 31.1% |
|  | 0.086 |  | 0.149 |  | 0.003 |  | 0.002 |  | 0.013 |  |
| Current tobacco use, based on the cotinine/ nicotine blood test (COT200) | - | 22.0% | - | 11.6% | - | 9.4% | - | 14.9% |  | 13.6% |
| Smoke tobacco use | 14.1% | 16.3% | 6.5% | 5.5% | 6.8% | 6.3% | 11.6% | 8.1% | 9.6% | 8.3% |
|  | 0.222 |  | 0.335 |  | 0.650 |  | 0.011 |  | 0.046 |  |
| All forms of current tobacco | 19.6% | 19.3% | 7.0% | 6.7% | 7.5% | 8.3% | 12.0% | 9.1% | 11.2% | 10.1% |
|  | 0.879 |  | 0.785 |  | 0.506 |  | 0.041 |  | 0.119 |  |
| Less than 5 servings of fruit and veg/ day | 87.7% | 85.2% | 91.7% | 83.8% | 75.1% | 82.5% | 94.9% | 94.4% | 87.8% | 86.4% |
|  | 0.146 |  | <0.001 |  | <0.001 |  | 0.630 |  | 0.067 |  |
| Inadequate physical activity levels | 3.9% | 1.5% | 6.7% | 4.4% | 4.1% | 4.4% | 2.6% | 3.2% | 4.4% | 3.6% |
|  | 0.003 |  | 0.023 |  | 0.739 |  | 0.438 |  | 0.074 |  |
| Sedentariness | 27.5% | 30.9% | 33.5% | 38.9% | 29.8% | 32.3% | 16.3% | 25.1% | 26.6% | 31.9% |
|  | 0.136 |  | 0.009 |  | 0.226 |  | <0.001 |  | <0.001 |  |

**NB: Figures below the prevalence rates are p-values comparing the prevalences**

**Table 4:** Prevalence of the common NCD Risk Factors by Urbancity.

| Risk factor | Urban |  | Rural |  | Total |  |
| --- | --- | --- | --- | --- | --- | --- |
|  | 2014 | 2023 | 2014 | 2023 | 2014 | 2023 |
| High blood pressure | 26.1% | 28.9% | 23.9% | 22.6% | 24.3% | 24.5% |
|  | 0.177 |  | 0.242 |  | 0.838 |  |
| High blood glucose | 2.0% | 2.9% | 1.1% | 2.8% | 1.3% | 2.9% |
|  | 0.205 |  | <0.001 |  | <0.001 |  |
| Overweight and obesity | 32.7% | 33.7% | 16.3% | 19.7% | 19.4% | 23.9% |
|  | 0.648 |  | <0.001 |  | <0.001 |  |
| Current alcohol use | 22.8% | 28.2% | 29.9% | 32.4% | 28.5% | 31.1% |
|  | 0.008 |  | 0.039 |  | 0.013 |  |
| Current tobacco use, based on the cotinine/ nicotine blood test (COT200) | - | 10.2% | - | 15.1% | - | 13.6% |
| Current smoke tobacco use | 6.2% | 7.1% | 10.4% | 8.9% | 9.6% | 8.3% |
|  | 0.436 |  | 0.053 |  | 0.046 |  |
| Current use of all forms of current tobacco | 6.7% | 7.8% | 12.2% | 11.1% | 11.2% | 10.1% |
|  | 0.359 |  | 0.192 |  | 0.119 |  |
| Inadequate consumption of fruits and vegetables | 89.4% | 86.5% | 87.4% | 86.4% | 87.8% | 86.4% |
|  | 0.054 |  | 0.259 |  | 0.067 |  |
| Inadequate physical activity | 7.8% | 6.5% | 3.5% | 2.3% | 4.4% | 3.6% |
|  | 0.282 |  | 0.007 |  | 0.074 |  |
| Sedentariness | 34.6% | 34.4% | 24.7% | 30.7% | 26.6% | 31.9% |
|  | 0.928 |  | <0.001 |  | <0.001 |  |

**NB: Figures below the prevalence rates are p-values comparing the prevalences immediately above them**

There was an overall significant increase in the prevalence of high blood glucose from 1.3% in 2014, to 2.9% in 2023 ((p< 0.001). Significant increases between the 2014 survey and the 2023 survey were observed in all regions of the country, except in the Northern region (p=0.735) - Table 3. Significant increases in the prevalence of high blood glucose were also observed in rural areas from 1.1% in 2014 to 2.8% in 2023 (p< 0.001). No significant changes were observed in urban areas between the two surveys – Table 4.

There was an overall significant increase in the prevalence of overweight and obesity, from 19.4% in 2014 to 24.1% in 2023 (p< 0.001). The significant increase was observed in three regions of the country, except the Eastern region (p= 0.163). There was also a significant increase the prevalence of overweight and obesity in rural areas from 16.3% in 2014 to 19.9% in 2023 (p< 0.001). No significant changes were observed in urban areas.

The prevalence of current alcohol consumption increased significantly from 28.5% in 2014, to 31.1% in 2023 (p=0.013). The significant increases occurred in all regions of the country except the Central region (p= 0.149) – Table 3. Significant increases in the prevalence of current alcohol consumption were also observed in both rural and rural areas between the 2014 survey and the 2023 survey (Table 4).

There was an overall significant degree in the prevalence of current smoke tobacco use from 9.6% in 2014, to8.3% in 2023 (p= 0.046). This significant decrease occurred in the Western region of the country only from11.6% in 2014, to 8.1% in 2023 (p= 0.011), Table 3. No significant differences were observed in the prevalence of smoke tobacco use in urban, or rural areas (Table 4).

Regarding current use of all forms of tobacco, overall, no significant differences between 2014 and 2023, only a marginal significant decrease was observed in the Western region from 12.0% in 2014, to 9.1% in 2023 (p= 0.041). No significant differences were observed in the prevalence of all forms of tobacco use in urban, or in rural areas (Table 4).

The overall prevalence of a positive cotinine/ nicotine test was 13.6%. This test was conducted during the 2023 survey only.

Overall, there was no change in the prevalence of inadequate consumption of fruits and vegetables, at 87.8% in 2014 and 86.4% in 2023 (p= 0.067). However, we observed a significant decrease in the prevalence of inadequate consumption of fruits and vegetables in the Eastern region from 91.7% in 2014, to 83.8% in 2023 (p< 0.001); whereas a significant decrease was observed in the Eastern region from 75.1% in 2014 to 82.5% in 2023. No significant changes were observed in the prevalence of inadequate consumption of fruits and vegetables in urban, or in rural areas (Table 4).

The prevalence of inadequate physical activity remained low at 4.4% in 2014 and 3.6% in 2023, but overall there were no significant changes between the two surveys (p= 0.074). However, we observed significant decrease between the two surveys in the prevalence of inadequate physical activity the Northern region from 3.9% in 2014 to 1.5% in 2023 (p= 0.003), and in the Central region from 6.7% I 2014 to 4.4% in 2023 (p= 0.023) (Table 3). A significant decrease in the prevalence of inadequate physical activity was also observed in rural areas from 3.5% in 2014 to 2.3% in 2023 (p=0.007) (Table 4).

The prevalence of sedentariness significantly increased overall from 26.6% in 2014, to 31.9% in 2023 (p< 0.001). The significant increases occurred in all regions of the country except the Eastern region (p= 0.226) – Table 3. Significant increases were also observed in rural areas, from 24.7% 2014, to 31.9% 2023 survey (p< 0.001) (Table 4).

Figures 1a and 1b show the overall prevalence of the NCD risk factors, comparing 2014 and 2023.

**Figure 1a:**
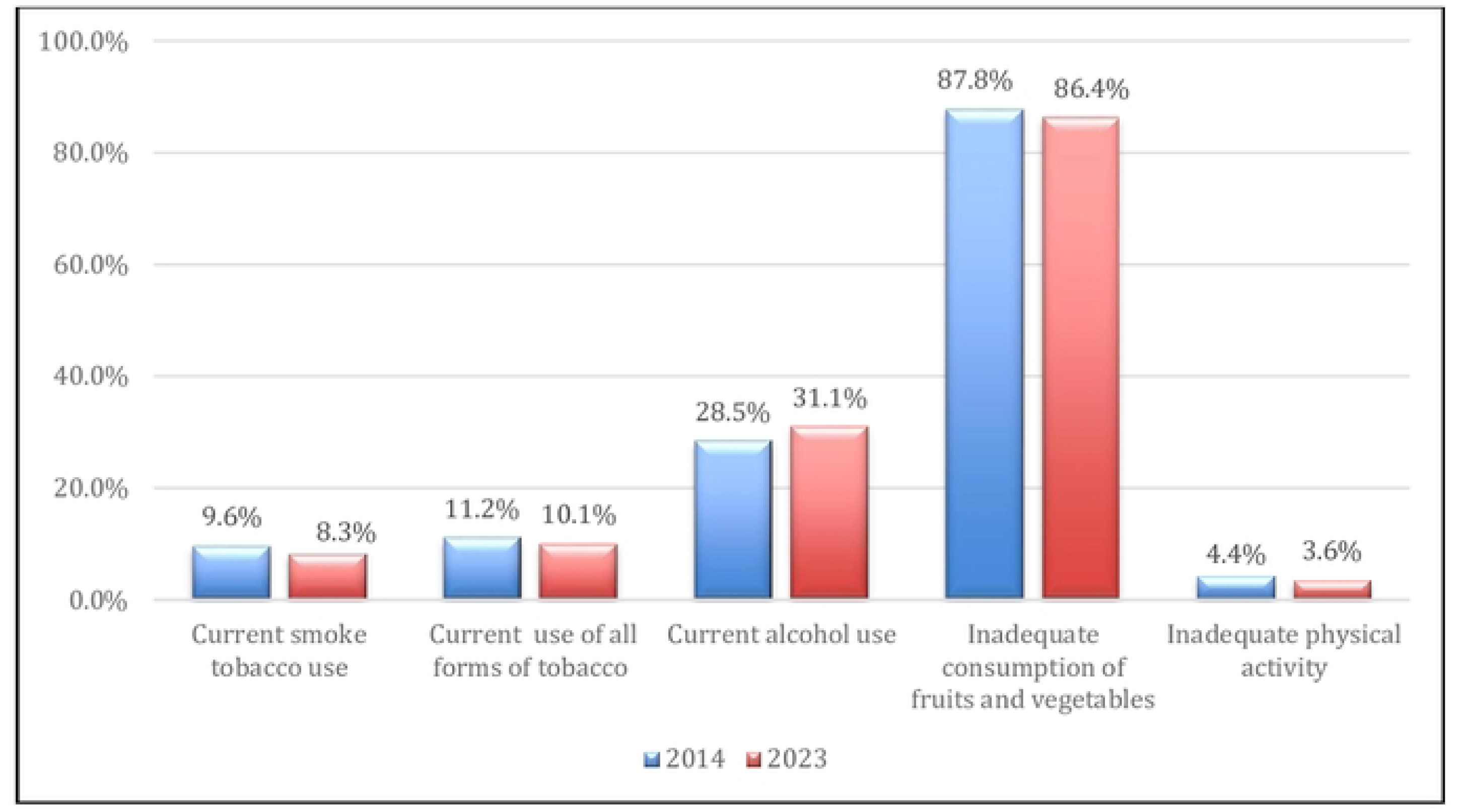
Prevalence of behavioral risk factors in 2014 and in 2023

**Figure 1b:**
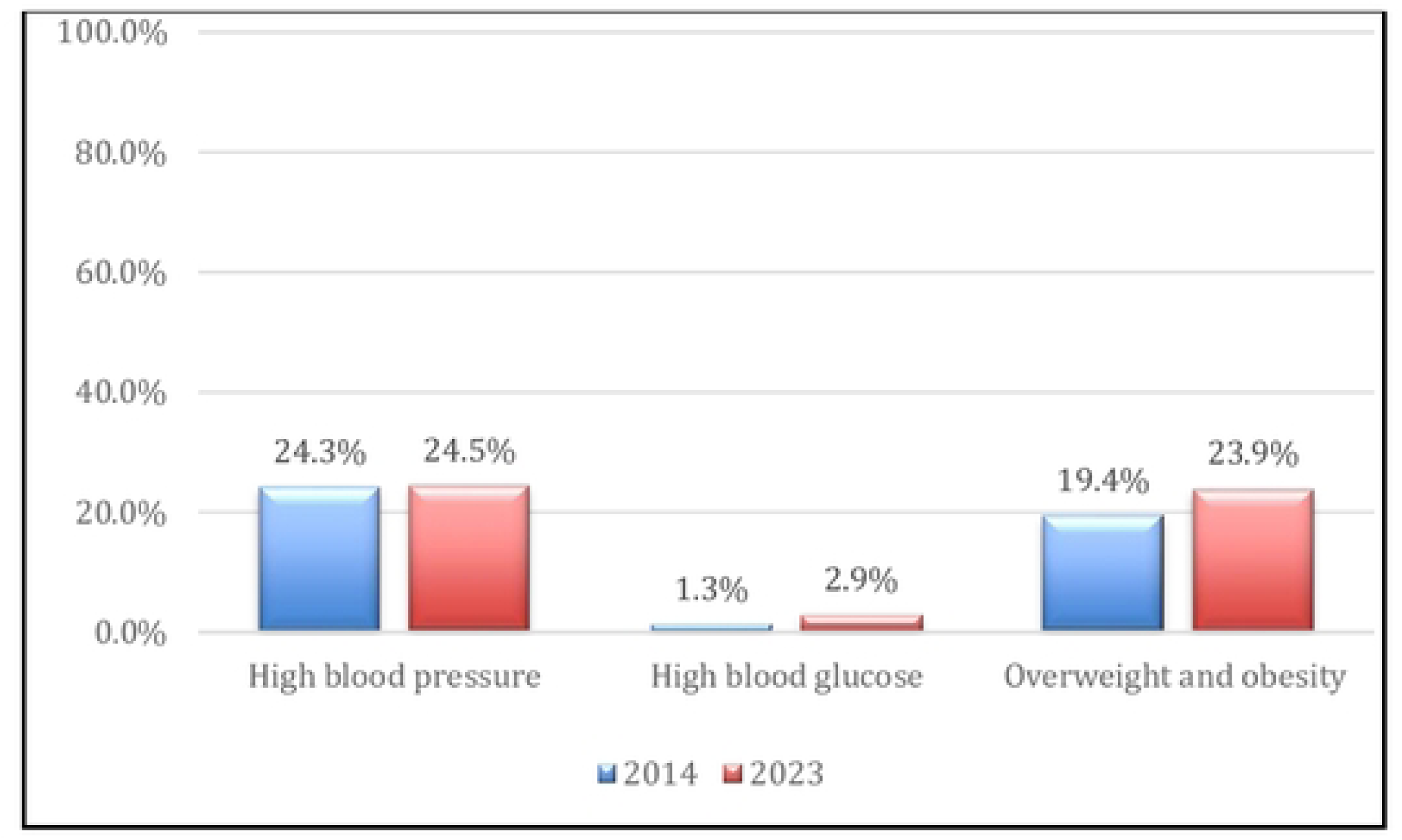
Prevalence of physiological /metabolic risk factors in 2014 and in 2023

## DISCUSSION

Our analysis reveals two important findings. First, the prevalence of most of the common NCD risk factors increased between 2014 and 2023, including: prevalence of high blood pressure, prevalence of overweight and obesity, prevalence of current alcohol consumption, and prevalence of sedentariness. Second, the prevalence of some of the risk factors remains unacceptably high including the prevalence of high blood pressure, being higher than 25%, and the prevalence inadequate consumption of fruits and vegetables, being higher than 80%. There is an agent need for the various stakeholders, including Uganda’s Ministry of Health, to identify and implement interventions aimed at reducing the prevalence NCD risk factors to prevent the currently increasing burden of NCDs and associated mortality that has been noted by the WHO Africa Regional Office [2]. NCDs threaten progress towards the 2030 Agenda for Sustainable Development Goals (SDG Target 3.4), which aim at reducing by one third by 2030 premature mortality from NCDs through prevention, treatment and promotion of mental health and well-being [16]. Despite the availability of cost-effective and evidence-based practices to address NCDs in LMICs [17], implementing these interventions remains a significant challenge, particularly in promoting healthy behaviors [18]. Thus concerted effort is required.

### Strengths and Limitations

An important strength of our analysis is that it is based on data collected from nationally representative sample surveys. Thus the prevalences reported represent a national picture. Second, the two surveys used the standardized WHO STEPS methodology, a methodology that has been validated and used in many LMIC countries, which makes the data reliable and findings comparable.

A limitation to our analysis is in regard to the self-reported behavioral NCD risk factors including current alcohol consumption, current tobacco use, physical inactivity, sedentariness. Self-report of behavioral characteristics is likely to have some recall bias, a form information bias which may lead to either over or under-estimation, and it is always difficult to determine the direction of bias, if any.

## Author Contributions

The authors contributed in the following ways: *Conceptualization:* SKB, DG & GNM; *Data curation:* RKusolo; *Formal Analysis:* MM, RKusolo & DG; *Funding Acquisition:* DG, GNM & SBK; *Investigation:* DG, SKB, GNM, RKusolo, RKajura & RW; *Methodology:* DG, SKB, RKusolo, RW; *Project Administration:* DG & SKB; *Resources:* DG & SKB; Software: RKusolo; *Supervision:* DG & SKB; *Validation:* DG, SKB, RW & RKusolo; *Visualization:* MM, RKusolo & DG; *Writing – Original Draft Preparation:* DG, RKusolo & MM; *Writing – Review & Editing:* DG, RKusolo, MM, SKB, RW, GNM & RKajura.

## Data Availability

The data on which analysis presented in this article can be accessed on request to Uganda's Ministry of Heath, through the corresponding author.

## Acknowledgements

The authors are grateful to the research participants who volunteered to participate in the NCD risk factors survey; and for the technical and administrative support provided by Uganda’s Ministry of Health, and that from the Uganda Bureau of Statistics. The authors also acknowledge the administrative support provided by the WHO Uganda Country Office through Hafisa Kasule and Christine Joan Karamagi. We also grateful for technical support provided by: 1) Cheick Bady Diallo, Regional Advisor-Strategic Information and Surveillance, NCD Communicable and Non-communicable Diseases Cluster (UCN), WHO Regional Office for Africa, Cité Djoué, Brazzaville, Congo; 2) Stefan Savin, Technical Officer, Surveillance, Monitoring and Reporting, Non-communicable Diseases Department, World Health Organization, Geneva, Switzerland; and 3) Patricia Rarau, Technical Officer, Surveillance, Monitoring and Reporting Non-communicable Diseases Department, World Health Organization, Geneva, Switzerland. The conduct of the two surveys on which findings reported in this article are based was supported with funding from: the Uganda government, the World Health Organization, the World Diabetes Foundation, and, the United Nations Development Program.

